# A Rapid Saliva Test for Monitoring Immune Protection against SARS -CoV-2 and its Variants

**DOI:** 10.1101/2021.07.09.21260224

**Authors:** Victoria SA. Momyer, Samantha Dixon, Q. John Liu, Brian Wee, Tiffany Y. Chen, Anna Petropoulos, L. Jessica Sang

## Abstract

Given the ongoing transmission and emergence of SARS-Cov-2 variants globally, it is critical to have a timely assessment on individuals’ immune responses as well as population immunity. Important questions such as the durability of COVID-19 immunity or the efficacy of vaccines require large datasets to generate meaningful insights. However, due to the complexity and relatively high-cost of many immunity assays and the needs for blood-drawing specialists, these assays were mostly limited to small-scale clinical studies. Our work demonstrated the potential of a non-invasive, inexpensive and data-driven solution for large-scale immunity surveillance and for predictive modeling of vaccine efficacy. Combining a proprietary saliva processing method and an ultra-sensitive digital detection technology, we were able to rapidly gather information regarding personalized immune response following infection or vaccination, monitor temporal evolution, and optimize predictive models for variant protection.

## INTRODUCTION

The COVID-19 pandemic, caused by infection with severe acute respiratory syndrome coronavirus 2 (SARS-CoV-2), has resulted in significant morbidity and mortality globally. Immunity to SARS-CoV-2 gained either through infection or vaccination has been shown to confer protection against reinfection and/or reduce the risk of clinically severe outcomes (*1-3*). However, the vaccination levels in many countries have not yet reached the threshold estimated to permit sufficient protection at the population level. There is ongoing transmission and emergence of viral variants that may escape control by humoral immune response induced by existing vaccines. For example, whereas the messenger RNA vaccine BNT162b2 (Pfizer–BioNTech) was shown to have an 89.5% efficacy in preventing disease caused by the B.1.1.7 variant, its efficacy against the B.1.351 infection was only 75% (*4,5*). Another challenge arose from the reported reduction in vaccine efficacy in elderly or immunocompromised patients (e.g. cancer patients receiving treatment) (*1,6-9*). It is not clear whether the duration of protective immunity is reduced in the immunocompromised patients or when a booster dose would be needed.

One of the gold standards to study immunity is through the measurements of neutralizing antibodies (nAbs) in serum (*10,11*). Measuring nAb has provided a deeper insight into understanding the vaccine efficacy on various subpopulations (*1,2*). It was also reported that nAb levels are highly predictive of immune protection from symptomatic SARS-CoV-2 infection (*3,10,11*). Most of the existing SARS-CoV-2 nAb assays were conducted in high-complexity labs using venous blood, which requires blood drawing by a trained phlebotomist (*12*). The commercially available SARS-CoV-2 antibody tests based on finger-prick blood mainly provide qualitative results and the test accuracy has been under scrutiny (*13*). Saliva, on the other hand, may serve as a simple non-invasive alternative to venous blood to monitor humoral immune responses on a large scale following SARS-CoV-2 infection or vaccination. Recent studies have indeed demonstrated that serum levels of IgG against Spike and receptor binding domain (RBD) positively correlate with IgG levels in matched saliva samples from convalescent patients, and also tightly correlate with the neutralizing titer (*7,14-16*). It is promising that the immune protection against SARS-CoV-2 and its variants can be estimated from a simple saliva test.

In the present study, we showcase TiMES (technology integrated magneto-electronic sensing), a novel digital technology platform for rapidly monitoring the neutralizing antibody levels against SARS-CoV-2 in human saliva samples. The technology has been previously demonstrated to accurately detect cancer, transplant rejection and sepsis from blood or urine specimens (*17-20*). It outperformed conventional assays in several aspects: the detection sensitivity was >100 fold higher compared to that of ELISA; the assay was fast, inexpensive and can be easily performed at the point of care (*17*). With the newly-developed TiMES-Now saliva assay, we were able to rapidly asses SARS-CoV-2 immunity following vaccination, monitor neutralizing antibody levels over time, and model immune protection against major variants. The test also allowed a closer examination on how different subpopulations, particularly immunocompromised patients, responded to the vaccines. TiMES-Now provides an affordable, data-driven and highly-accurate solution for the rapid assessment of personalized immune response as well as population immunity, and to inform vaccination strategy.

## RESULTS

### A rapid and automated TiMES-Now assay for detecting SARS-CoV-2 neutralizing antibodies (nAb) in saliva

In order to monitor the neutralizing antibody response to SARS-CoV-2, we developed the rapid and automated TiMES-Now nAb assay based on integrated magneto-electronic sensing (iMES). We initially focused on the IgG antibody to the Spike RBD, as neutralizing antibodies are directed to the Spike protein and the RBD domain plays key roles in viral entry. The assay incorporates a dual signal-amplification scheme to achieve a superior detection sensitivity to existing rapid antibody tests. The assay first uses magnetic particles (MP) to capture antibodies against the SARS-Cov-2 RBD protein directly from saliva and subsequently labels them with enzyme-linked anti-human IgG antibodies. MP-antibody complexes are then magnetically concentrated on top of a sensing electrode; redox reactions and electron transfer from the electrode generated electrical current as an analytical readout (**Fig. 1a**). Both magnetic concentration and enzymatic reaction serve to amplify the analytical signal, boosting the detection sensitivity.

**Fig. 1.**
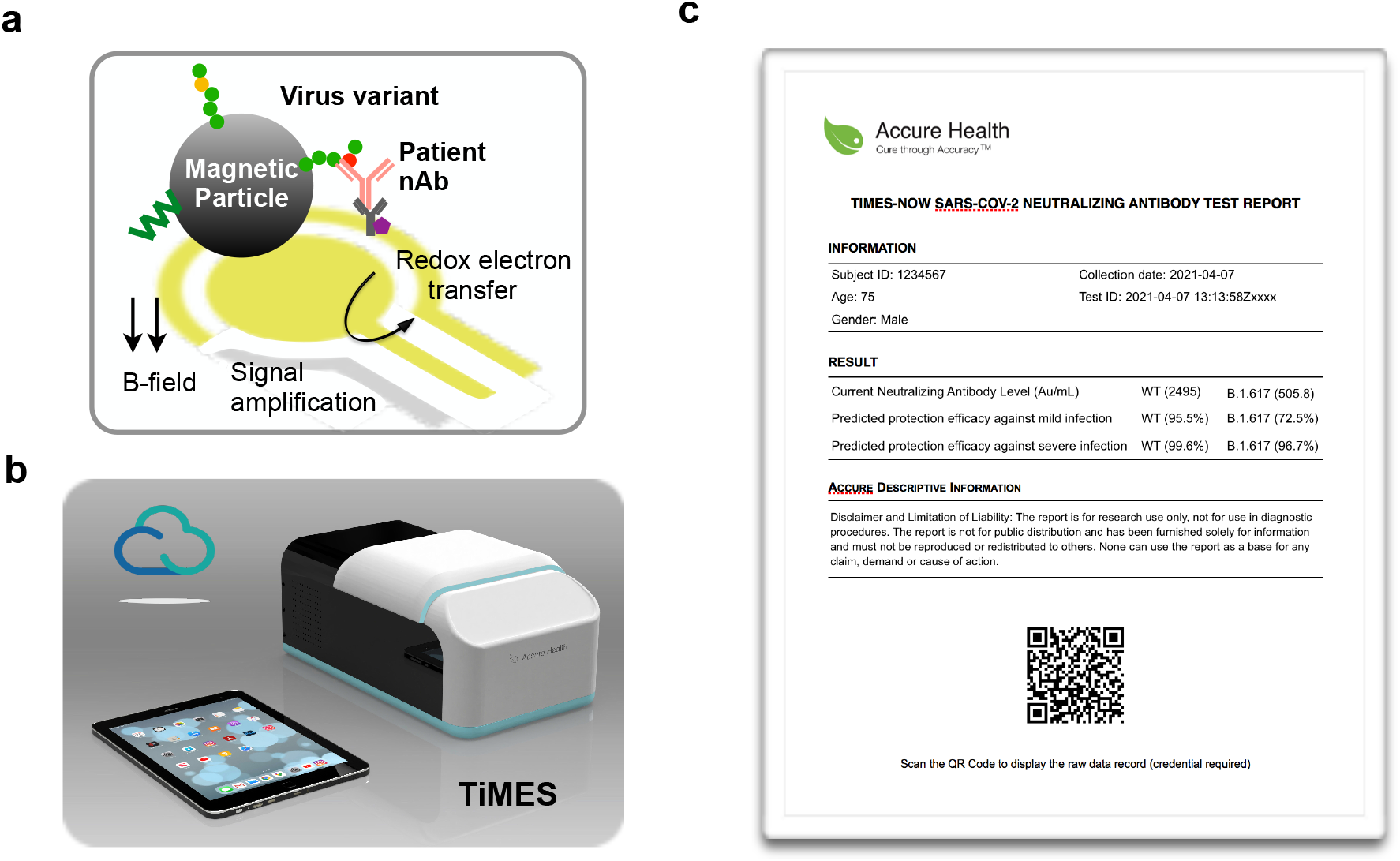
TiMES-Now SARS-Cov-2 nAb assay schematic and TiMES automation system. **(a)** IgG antibodies against the SARS-Cov-2 RBD protein were captured on magnetic particles directly from saliva and labeled with secondary antibodies for electronic detection. **(b)** TiMES is a compact, connected automation system controlled wirelessly by a mobile tablet. Up to 8 tests can be performed in parallel and completed in 30 min. **(c)** Once the test is done, the mobile APP conducts automatic analyses and provides a test report in real time. Both the measured neutralizing antibody level and the estimated immune protection efficacy are provided in the report.

To improve throughput and reproducibility of the SARS-Cov-2 nAb assay, the tests were performed using a portable and automated TiMES system (Accure Health). The newly developed TiMES is a next-generation system advanced from earlier prototypes of the iMES device (*17-19*). The TiMES system features i) automated assay process; ii) 8x parallel detections; and iii) real-time data analyses and secure cloud integration. Robotic design, electronics, and firmware were optimized to achieve “sample-in-answer-out” workflow automation. TiMES has a small footprint (∼10’’ × 7’’ × 6’’; 26 cm x 18 cm x 15 cm) and can be easily set up in an office or home setting (**Fig. 1b**). Proprietary test cartridges and mobile TiMES APP were also customized for conducting the TiMES-Now SARS-Cov-2 nAb tests. Up to 8 tests can be performed in parallel and completed in 30 min. Once the assays are completed, the mobile TiMES APP conducts automatic analyses and provides individual test reports in real time (**Fig. 1c, Table 1**).

**Table 1.** TiMES-Now SARS-Cov-2 nAb Assay.

|  |  |  |
| --- | --- | --- |
| **Validated sample type** |  | Saliva, serum |
| **Analyte type** |  | SARS-Cov-2 Spike RBD IgG antibodies, including variants |
| **Analytical performance** | Reproducibility | CV< 5% |
|  | Dynamic range | > 4 orders of magnitude |
| **Test throughput** |  | 8 tests/device/30 min (automated test and data report), hands-on time ~5min |
| **Data analysis** |  | Mobile APP for real-time analysis and reporting, sync with cloud and HIPAA-ready |

For quantitative analyses, we established a standard curve for each production batch, and integrated the standard curve as a built-in function in the TiMES APP. The range of the standard curve was selected based on physiologically-relevant concentrations of anti-SARS-Cov-2 Spike IgG antibodies found in patient samples. Varying concentrations of anti-SARS-Cov-2 Spike IgG antibodies were diluted into a negative saliva sample (no history of COVID-19 exposure or vaccination) and assayed. The TiMES-Now anti-SARS-Cov-2 nAb assay achieved excellent fitting (R^2^ = 0.998) in the selected analytical range and reached a limit of detection of ∼5 pg/mL (**Fig. S1**).

### TiMES-Now enables SARS-CoV-2 nAb monitoring from convalescent and vaccinated patients

To ensure accurate and reproducible results, we investigated the consistency of the TiMES-Now SARS-CoV-2 nAb assay. Non-stimulated saliva samples were self-collected from study participants who were fully vaccinated (Materials and Methods). Aliquots of saliva samples were prepared, stored at 4 °C and tested within 48 hrs of collection. Coefficient of variation (CV = s.d./mean) was calculated for each set of aliquot measurements. TiMES-Now SARS-CoV-2 nAb assay achieved high reproducibility, with a CV < 5% (**Fig. S2a**). We further studied whether saliva collection time affects the test results. Three independent saliva samples were collected from the same study participant within a 24 hr time frame (evening, morning, afternoon) and tested. TiMES-Now SARS-CoV-2 nAb assay achieved a CV < 7%. Hydration levels and circadian rhythms may have contributed to the test variation but the overall reproducibility was still acceptable (**Fig. S2b**). We also tested if beverages or oral hygiene products affect the test results. A small amount of coffee or toothpaste solution were added to the saliva aliquots and compared with control aliquots diluted with distilled water. Both coffee and toothpaste resulted in a reduction in signals from TiMES-Now SARS-CoV-2 nAb assay (**Fig. S2c**). It was therefore recommended that the study participants rinse their mouth with water prior to the saliva self-collection.

We evaluated the performance of the TiMES-Now SARS-CoV-2 nAb assay using freshly-collected saliva samples from convalescent or vaccinated patients (Materials and Methods; **Table 2**). Compared with samples from study participants who were neither infected nor vaccinated (n=45), samples from COVID-19 subjects were mostly positive for SARS-CoV-2 nAb (n=10; AUC=0.92) between 1-9 months post infection (**Fig. 2a**). The majority of saliva samples from patients that were tested at least 15 days after receiving a COVID-19 vaccine (Pfizer, Moderna, J&J, Sinopharm, Novavax) also showed positive SARS-CoV-2 nAb results (n=67; AUC=0.97) (**Fig. 2b**).

**Table 2.**
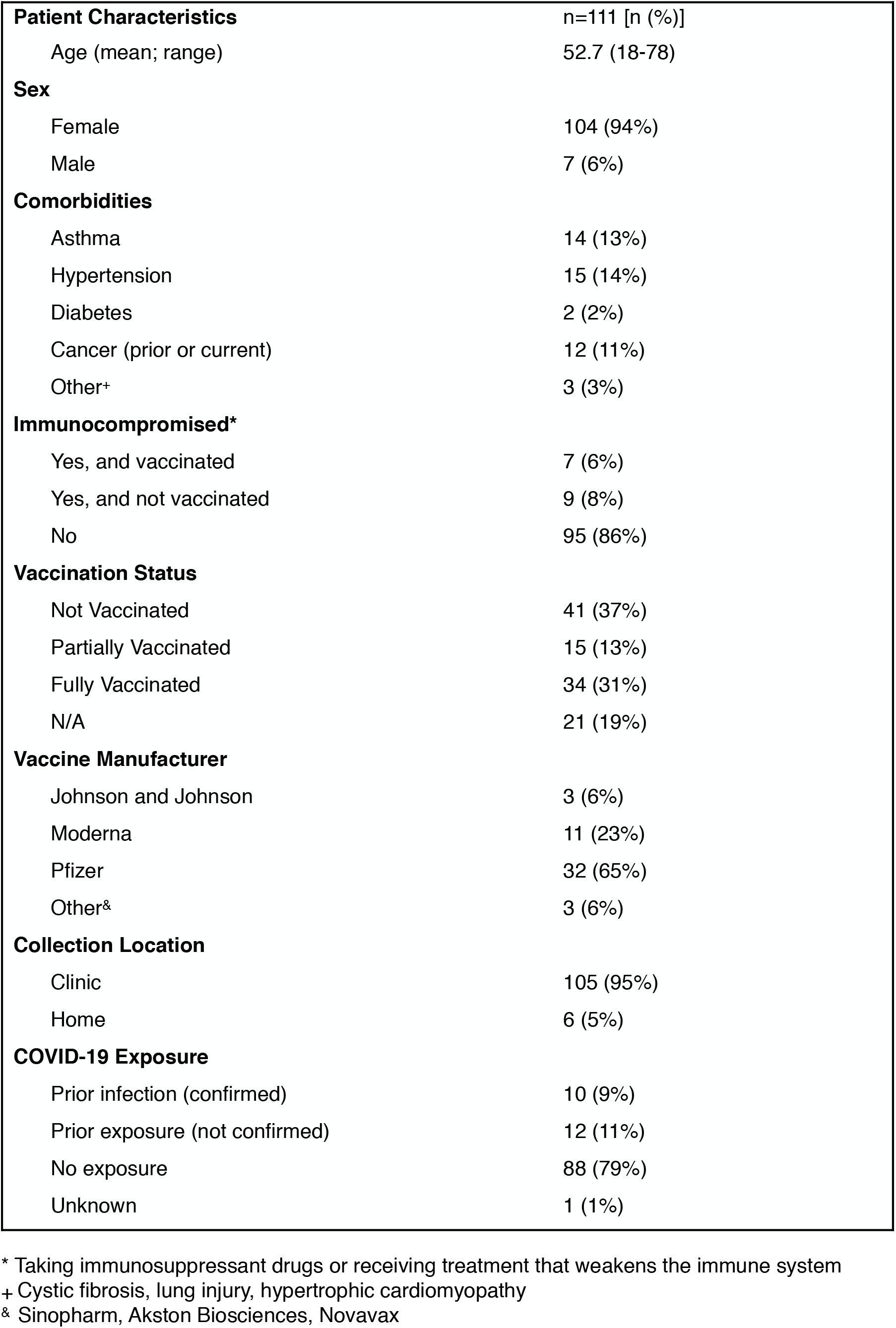
Patient Demographics.

**Fig. 2.**
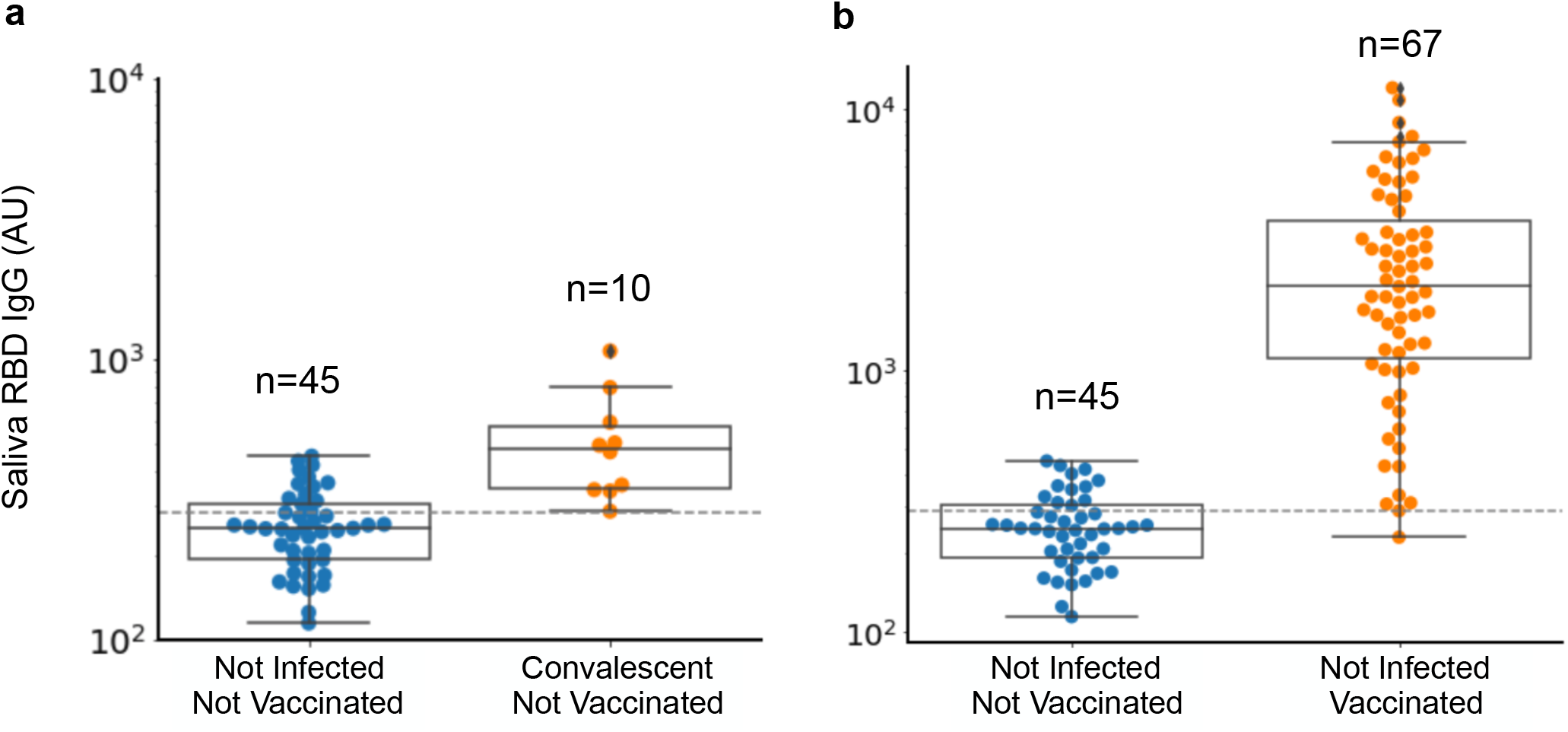
TiMES-Now enables SARS-CoV-2 nAb monitoring from convalescent and vaccinated patients. Saliva samples were collected from participants who were neither infected nor vaccinated (n=45), and from **(a)** convalescent participants who were previously tested positive for COVID-19 with a PCR test and not yet vaccinated (n=10), and **(b)** vaccinated participants who were previously immune naive and at least 15 days post vaccination (n=67). ROC curves were generated and the areas under the curve (AUC) were calculated. A cutoff level was selected and shown in dotted lines. The saliva SARS-CoV-2 nAb levels were induced in **(a)** convalescent and **(b)** vaccinated participants. Boxes and horizontal bars denote interquartile range (IQR) and median value, respectively. Whisker endpoints are equal to the maximum and minimum values below or above the median ±1.5 times the IQR. The data are presented on a logarithmic scale.

To better understand the temporal evolution of immune response, we conducted longitudinal monitoring of the nAb levels using the TiMES-Now SARS-CoV-2 saliva nAb assay. The test was able to detect positive SARS-CoV-2 nAb from saliva about one week after the participants received the first dose of a COVID-19 mRNA vaccine. The saliva nAb levels peaked after the second dose, stayed high over a period of ∼120 days, and then started to decline around ∼120 days post vaccination (**Fig. 3**). These results were consistent with the published nAb titers from serum samples (*21,22*).

**Fig. 3.**
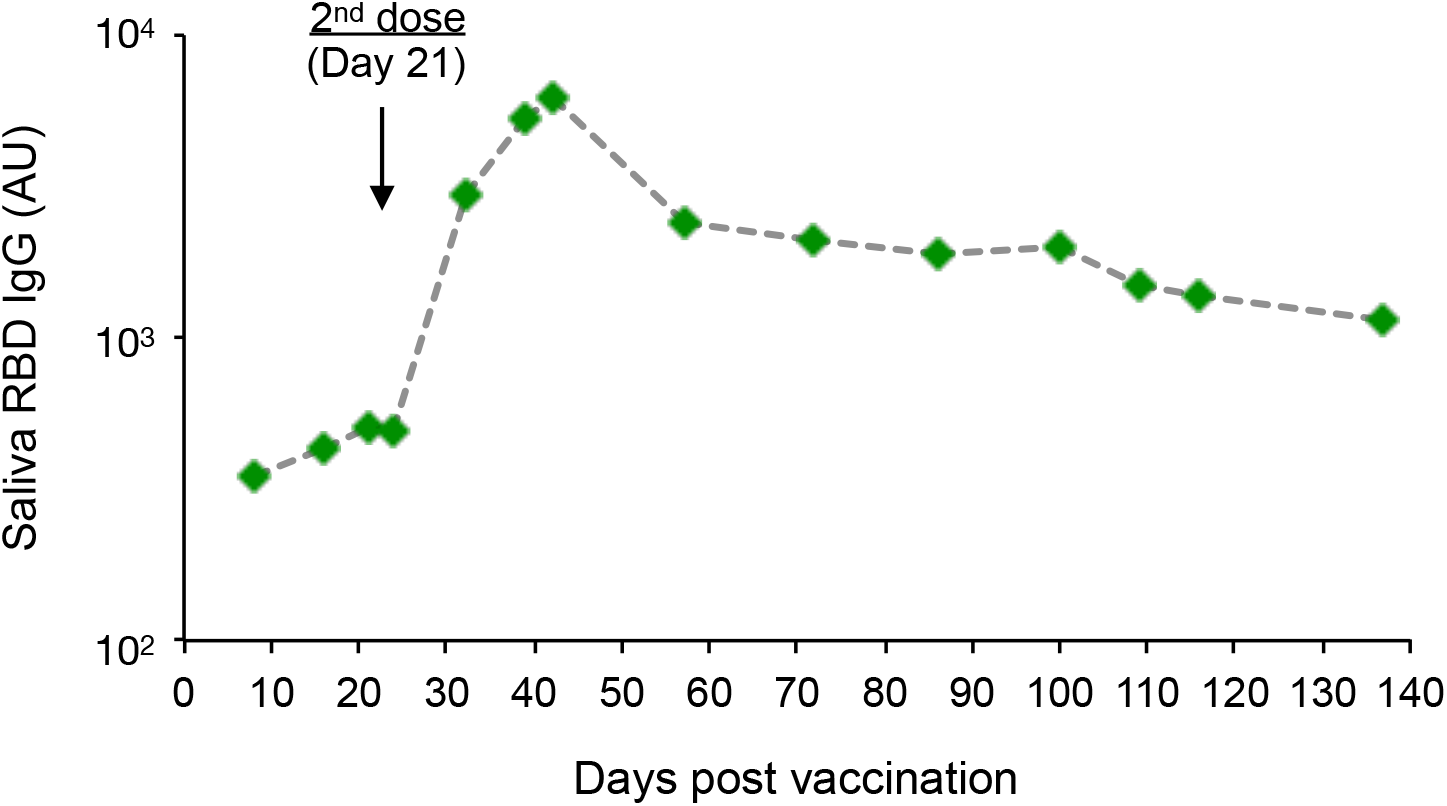
Longitudinal monitoring of SARS-Cov-2 RBD IgG responses in saliva. Saliva samples were collected from an immunologically naive study participant at the indicated time points after vaccination (3∼14 days interval). Sample were tested with the TiMES-Now SARS-Cov-2 saliva nAb assay and the TiMES readings were plotted and connected with a dotted line. TiMES-now was able to detect positive SARS-CoV-2 nAb from saliva one week post-vaccination. The nAb levels showed steady increases after the first vaccine dose, and peaked about two weeks after the second dose. The data are presented in the logarithmic scale.

Recent studies suggest that neutralizing antibody levels are highly predictive of immune protection from symptomatic SARS-CoV-2 infection (*10,11*). We thus analyzed the saliva nAb levels from some “high-risk” sub-populations and estimated whether there were sufficient nAb for immune protection.

Consistent with the recently published studies on vaccine efficacy (*6,7*), we observed that immunocompromised individuals (e.g. patients taking immunosuppressant drugs or receiving treatments which may weaken the immune system) had relatively lower levels of nAb than healthy participants (**Fig. 4a**). While the second dose generally provided a boost to the nAb levels, the overall levels of nAbs from immunocompromised patients were still lower than the healthy controls (**Fig. 4b-c**). We estimated the saliva nAb level for 50% protection against detectable SARS-CoV-2 infection based on a recently published predictive model (*11*). At the time of testing, the nAb levels from these immunocompromised patients were still above the 50% efficacy threshold. However, modeling of the nAb decay over the first 120 d after immunization suggests that a significant loss in protection from SARS-CoV-2 infection may occur sooner in these “high-risk” patients.

**Fig. 4.**
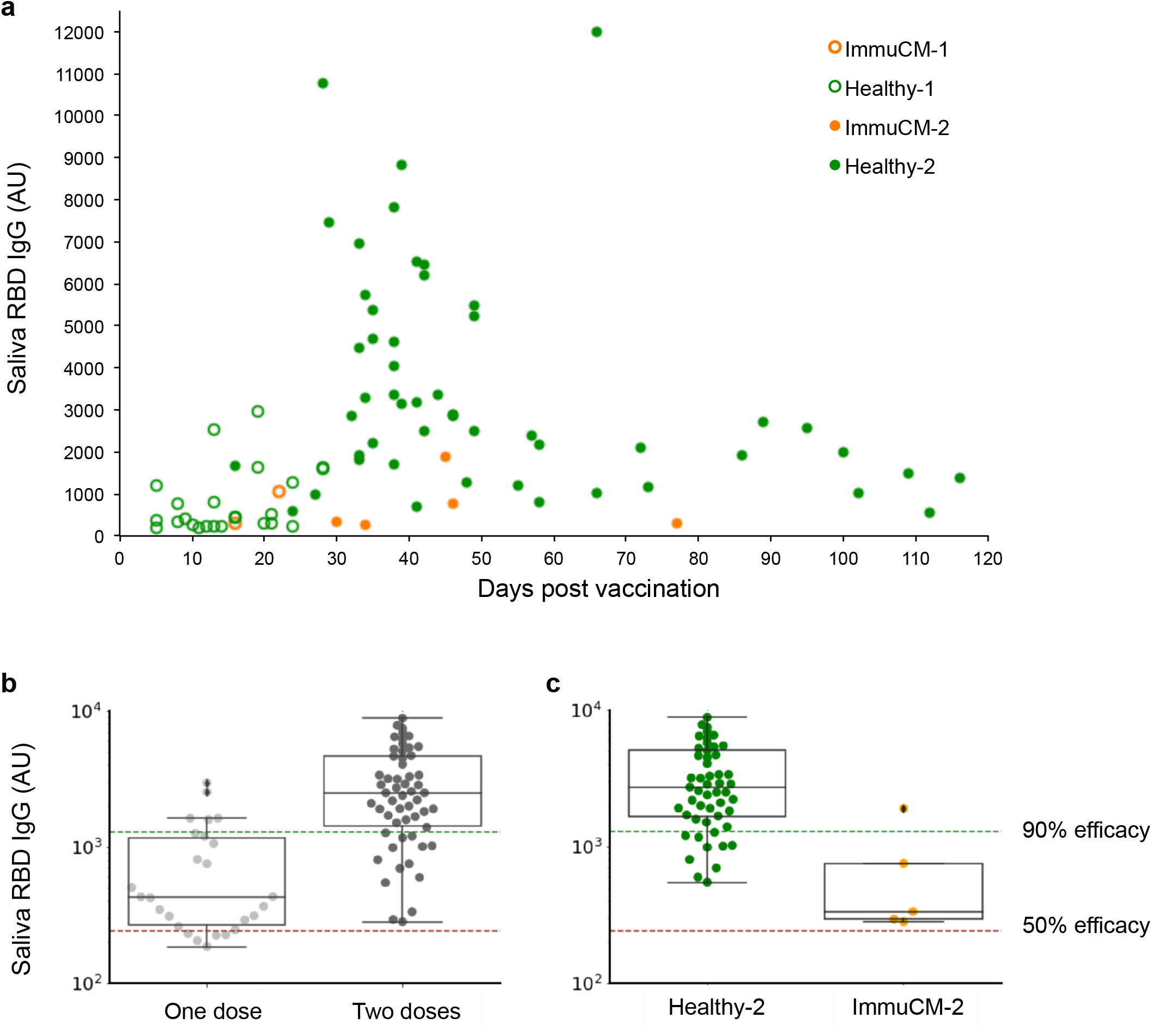
Longitudinal and dose-dependent analysis of SARS-Cov-2 RBD IgG responses in immunocompromised patients. **(a)** At the indicated time points after vaccination, saliva samples were collected from immunocompromised (ImmuCM) individuals and healthy participants (healthy). Sample were tested with the TiMES-Now SARS-Cov-2 saliva nAb assay and the TiMES readings were plotted. The overall SARS-Cov-2 nAb levels were much higher after study participants received both vaccine doses. **(c)** Even after the second vaccine dose, the levels of nAbs from immunocompromised patients (ImmuCM-2) were still lower than the healthy cohort (Healthy-2) (*p* < 0.002). Boxes and horizontal bars denote interquartile range (IQR) and median value, respectively. Whisker endpoints are equal to the maximum and minimum values below or above the median ±1.5 times the IQR. The predictive thresholds for reaching 50% and 90% protection efficacy are also indicated. The data are presented in the arithmetic (**a)** and the logarithmic **(b, c)** scales.

### TiMES-Now saliva assay monitors immune protection against viral variants

In addition to the declining neutralization titer over time, reduced vaccine efficacy to different viral variants have also been reported (4,*5*). Particularly, it has been shown that the neutralization titer against the B.1.351 variant in vaccinated individuals was significantly lower compared with the early wild-type strain (*4,5*). The gold standard for measuring the neutralization titer is based on venous serum samples. Recent studies, however, have demonstrated the potential use of saliva: IgG against Spike and RBD in the serum positively correlated with matched saliva samples from convalescent patients, and also tightly correlated with the neutralizing titer (*7,14-16*). We thus applied the TiMES-Now SARS-CoV-2 nAb assay to test whether there was a positive correlation between the saliva nAb levels and the serum nAb levels, to different viral variants. Matched saliva and serum samples from five fully-vaccinated patients were tested to measure the levels of nAb to the wild-type Spike RBD as well as to two of the variants of concern: B.1.1.7 and B.1.351. Our results showed a high correlation (R^2^=0.934) between the saliva nAb levels and the serum nAb levels, including their response to the variants (**Fig. 5a**). The data also supported a reduction in nAb efficacy against the B.1.351 variant. Taken together, the TiMES-Now SARS-Cov-2 nAb saliva assay demonstrated rapid and accurate assessment of variant response in vaccinated patients.

**Fig. 5.**
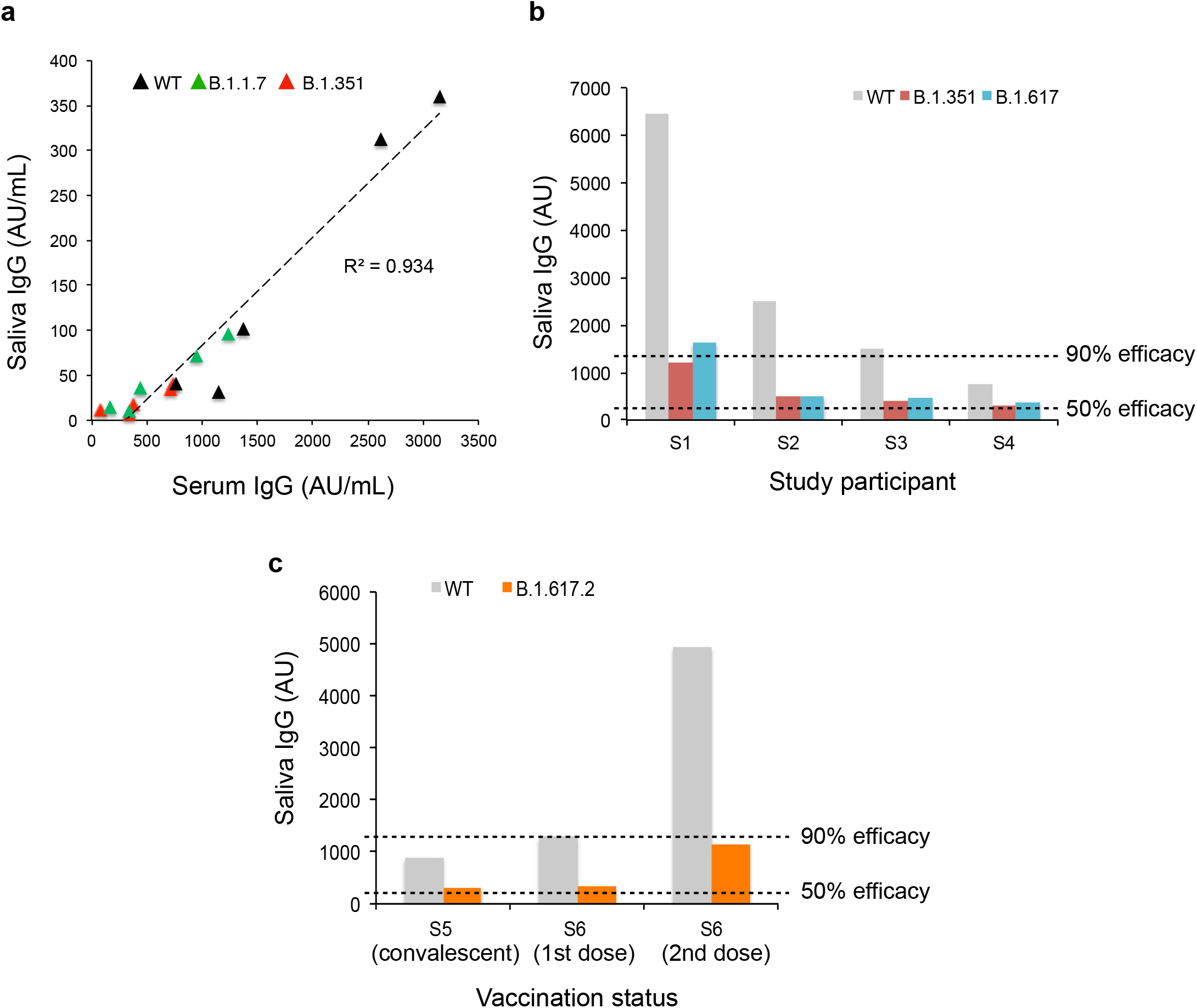
TiMES-Now saliva assay monitors the effect of viral variants on immune protection. **(a)** Correlation of RBD IgG responses in serum and saliva. Five matched serum and saliva sample pairs were tested using the TiMES-Now SARS-Cov-2 assay for correlations in levels of anti-Spike RBD IgG, including IgG to the wild-type viral antigen and two of the viral variants. The results showed high correlation between matched serum and saliva samples (*R*2 = 0.934). **(b)** The IgG responses toward viral variants (B.1.351 and B. 1.617) were tested using saliva from 4 vaccinated participants, including an immunocompromised patient S4. The predicted protection against B.1.351 and B1.617 were lower. Temporal responses toward B.1.617.2 (aka. Delta variant) were tested using saliva from a non-vaccinated convalescent participant S5, or from a vaccinated participant S6 following the 1st and the 2nd dose of a COVID mRNA vaccine. The protection against B.1.617.2 was enhanced by the 2nd dose. The predictive thresholds for reaching 50% and 90% protection efficacy are indicated.

Predictive modeling suggests that a lower neutralization titer is likely to reduce efficacy for immune protection (*11*). We therefore examined the nAb levels and estimated the efficacy for immune protection against several variants using the TiMES-Now SARS-Cov-2 saliva nAb assay. We focused on three emerging variants (B.1.351, B1.617, B1.617.2), as they harbor critical mutations which may enhance viral transmissions. While the predicted immune protection against the wild-type strain was mostly above the 90% efficacy threshold, the predicted protection against these variants were lower, particularly for an immunocompromised patient S4 (**Fig. 5b**). Two doses of COVID-19 mRNA vaccination enhanced the immune protection against B1.617.2 (aka. Delta variant)(**Fig. 5c**).

## DISCUSSION

Given the ongoing transmission and emergence of viral variants globally, it is critical to have a timely assessment on the personalized immune response as well as the population immunity, in order to better plan the next steps in the COVID-19 vaccine program. Immunity evaluation through measurements of neutralization antibody titer has been shown to be an important predictor of vaccine efficacy (*11,21-23*). However, due to the complexity and relatively high-cost of many conventional neutralization assays and the needs for blood-drawing by trained specialists, these assays were largely limited to small-scale clinical studies. Our work introduced a non-invasive, highly-accurate and inexpensive solution for large-scale immunity surveillance and for predictive modeling of vaccine efficacy. Combining a proprietary saliva processing formula and an ultra-sensitive digital detection technology, the TiMES-Now SARS-Cov-2 saliva nAb assay achieved highly-reproducible results in 30 min with a portable, digital system.

The current study demonstrated the potential of TiMES-Now SARS-Cov-2 saliva nAb assay to rapidly assess the induction of SARS-CoV-2 immunity in “high-risk” populations, monitor temporal response, and predict protection against viral variants. Some of the limitations of this study include a relatively small dataset, a predictive algorithm based on analyzing aggregated data from clinical trials and using a published but unverified data model (*11*). In the future, the predictive algorithm can be further improved with larger datasets, which can be readily collected with the TiMES digital platform.

Another limitation of the current study was to focus on immunity monitoring based on neutralizing antibodies alone. A number of studies have suggested that memory B cells and T cells may also contribute to long-term immunity and the protection against future infections (*23-26*). While the current study did not address B-cell or T-cell responses, the TiMES digital system has allowed us to rapidly develop a number of assays to investigate the immune response mediated by cytokines as well as other B-cell and T-cell biomarkers (*18,19, 27*). Together they will provide a more complete understanding of the SARS-CoV-2 immunity, inform vaccination strategy, and better prepare us for future outbreaks.

## MATERIALS AND METHODS

### Clinical Study

Study subjects were consented and enrolled in the study with approval from the Biomedical Research Alliance of New York Institutional Review Board (BRANY IRB # A20-08-597-840). Subject recruitment, and saliva collection were conducted at the New England Facial & Cosmetic Surgery Center in Danvers, Massachusetts. Saliva was self-collected at the study site and submitted to the study staff. In cases where the study subject was not able to visit the study site, saliva collection was self-administered at home and delivered to the study site. A screening questionnaire was completed by each study subject, and de-identified questionnaire data were collected and analyzed. Immunocompromised individuals were patients taking immunosuppressant drugs or receiving treatments which may weaken the immune system. Non-vaccinated convalescent patients were adults with a prior positive COVID-19 PCR test by self-report who met the definition of recovery by the Centers for Disease Control. Full cohort and demographic information is provided in **Table 2**.

### Sample collection and processing

Whole saliva was collected from a subject without any stimulation. A small amount of proprietary Saliva Processing Matrix (Accure Health, patent pending) was pre-loaded into the saliva collection tube to help improve sample safety and quality. After saliva collection, sample was mixed thoroughly with the Saliva Processing Matrix in a closed collection tube. Samples were tested within 48 hrs of collection, or aliquoted and stored at -80°C. For matched serum and saliva samples, venous blood was collected by standard phlebotomy into a BD serum tube. Serum was separated by centrifugation for 10 minutes at 2000g and then stored at -20C. Saliva was collected within 24hrs of blood collection.

### TiMES-Now SARS-Cov-2 nAb assay

40uL of saliva (or 10uL of serum) was loaded to the TiMES-Now SARS-Cov-2 nAb test cartridge (ISO 13485 certified), inserted into a TiMES automation device. Tests were performed using the TiMES system. Results were generated automatically with the TiMES APP and transferred to a password-protected, encrypted cloud database to meet HIPAA compliance. To establish a standard curve, varying concentrations of anti-SARS-Cov-2 Spike IgG antibodies (Sino Biological) were spiked into a negative saliva sample and assayed using the TiMES automation system. A curve was fitted using the TiMES readings and the antibody concentrations. The TiMES system achieved high intra-cartridge and inter-cartridge consistency from testing antibody standards. A built-in standard curve was therefore generated for each production batch, saved in the TiMES APP, and automatically calibrated to provide an accurate antibody concentration for each test. For variant studies, viral antigens were obtained from a commercial vendor (Sino Biological: 40591-V08H10, 40591-V08H12, 40592-V08H88, 40592-V08H90) and integrated into the TiMES-Now test cartridges.

### Modeling of the relationship between saliva nAb level and immune protection

To model the protective efficacy using the data from the TiMES-Now SARS-Cov-2 saliva nAb assay, we first established an equation to convert the TiMES-Now SARS-Cov-2 saliva nAb readings into corresponding serum neutralization titer, based on data from the present study and the published clinical trial data (*21*). Two assumptions were made: (i) the 50% geometric mean neutralization titer presented in Walsh 2020 (*21*) is equivalent to that in the present study; (ii) saliva and serum readings are highly correlated (R^2^ = 0.934). Once the TiMES-Now SARS-Cov-2 saliva nAb readings were converted to serum neutralizing titer, we adapted a predictive logistic model (*11*) to arrive at the protective efficacy against SARS-CoV-2 infection:

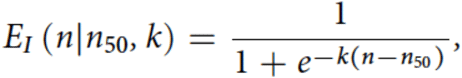

where *E*^*I*^ is the protective efficacy of an individual given the neutralization level *n*. The parameter *n*_50_ is the neutralization level at which an individual will have a 50% protective efficacy (that is, half the chance of being infected compared with an unvaccinated person). The steepness of this relationship between protective efficacy and neutralization level is determined by the parameter *k*. From the TiMES-Now SARS-Cov-2 saliva nAb level, we further estimated the time to reach a certain protective efficacy (50% or 70%) based on the published nAb half-life (*11*).

## Data Availability

The main data supporting the results in this study are available within the paper and its supplementary information. The raw patient datasets generated and analyzed during the study are available from the corresponding authors on reasonable request, subject to approval from the Biomedical Research Alliance of New York Institutional Review Board.

## ACKNOWLEDGEMENTS

The authors thank Jennifer Moran and staff at the New England Facial & Cosmetic Surgery Center for assisting with the research. We thank Dr. Karen Heichman and Dr. Daniel Wollin for helpful discussion and/or reviewing the manuscript.

## AUTHOR CONTRIBUTIONS

V. SA. Momyer, Q.J. Liu, T.Y. Chen, A.Petropoulos, and L.J. Sang designed the study, prepared figures, and wrote the manuscript. B. Wee contributed to the TiMES system development. V. SA. Momyer, S. Dixon and Q.J. Liu performed research and analyzed data. T.Y. Chen and A.Petropoulos supervised the clinical study.

## FIGURE LEGENDS

**Fig. S1.**
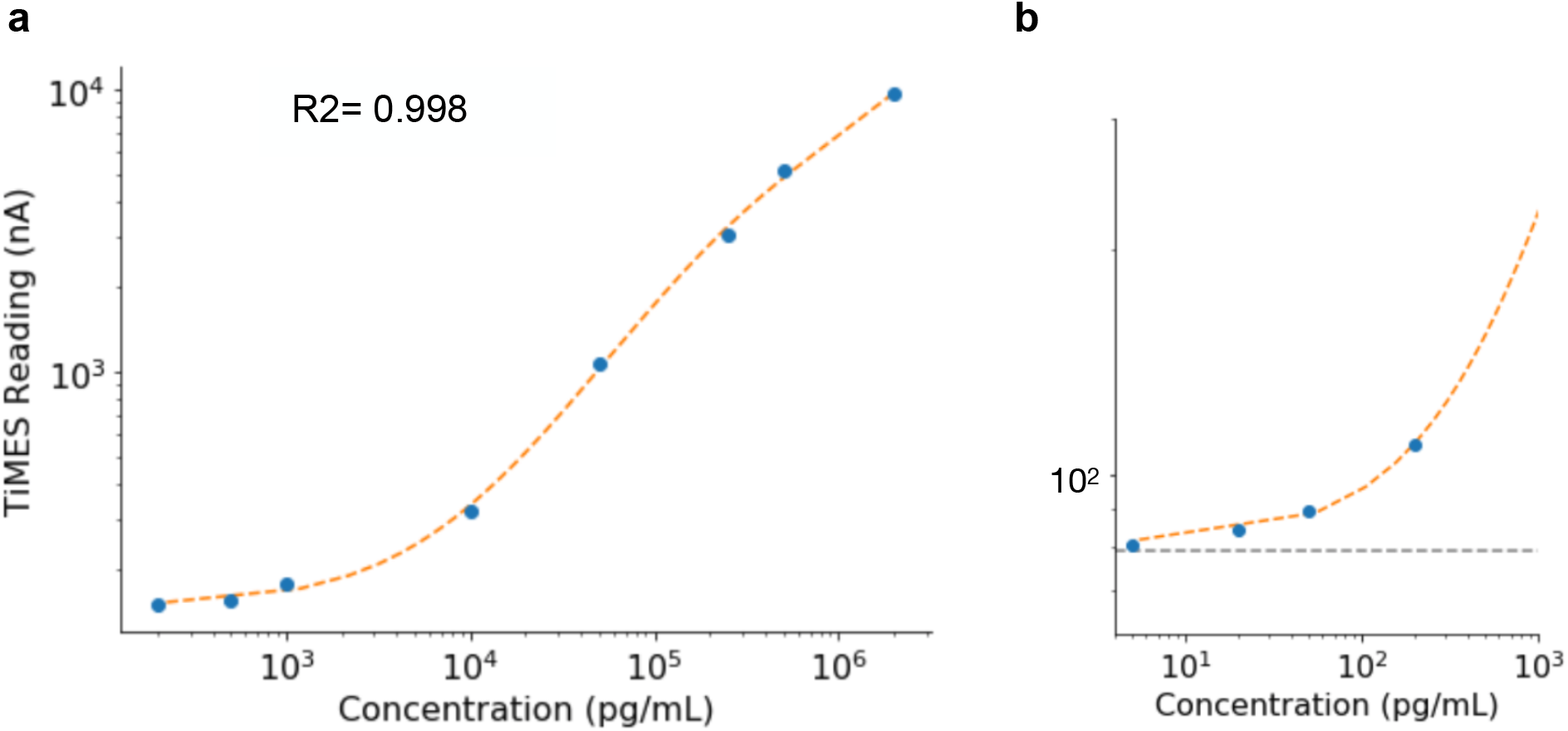
TiMES-Now SARS-Cov-2 nAb assay standard curve and limit of detection. **(a)** Varying concentrations of anti-SARS-Cov-2 Spike IgG antibodies were diluted into a negative saliva sample and assayed using the TiMES automation system. The TiMES readings were plotted and a standard curve was generated. The test achieved a large dynamic range, and a high correlation (R^2^ = 0.998) in the analytical range found in clinical samples. **(b)** TiMES achieved a limit of detection of ∼ 5 pg/mL. The data are presented on a logarithmic scale.

**Fig. S2.**
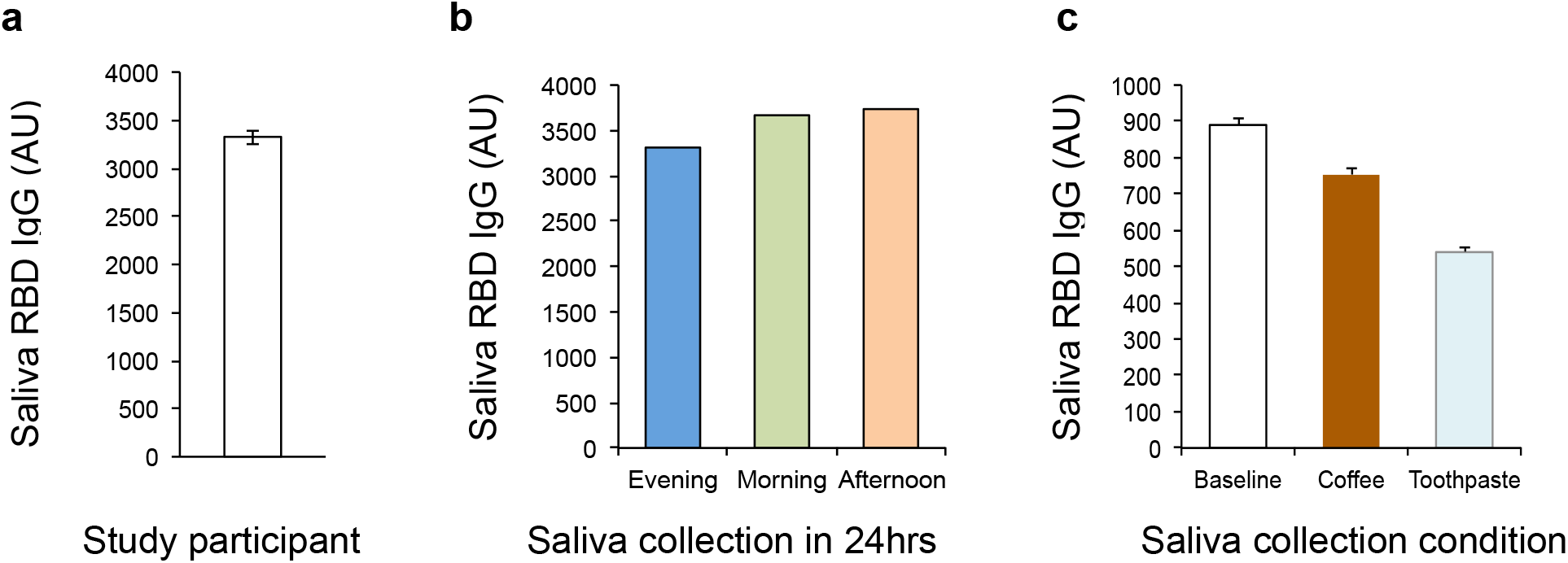
TiMES-Now SARS-Cov-2 nAb saliva assay achieved highly-reproducible results. **(a)** Non-stimulated saliva samples were self-collected from one study participant. Aliquots of saliva samples were prepared and tested within 24hrs of collection. There was high consistency between aliquot results. **(b)** Three independent saliva samples were collected from one study participant over a period of 24 hrs (evening, morning, afternoon) and tested. There was only small variation (CV< 7%) among test results. **(c)** A small amount of coffee or toothpaste solution were spiked into the saliva aliquots and compared with control aliquots diluted with water. Coffee and toothpaste may affect test results. All measurements in **(a)** and **(c)** were performed in duplicates, and the data are displayed as mean ± SD. AU: arbitrary units from the TiMES readings.

